# Interaction between risk SNPs of Developmental Dyslexia and Parental Education on Reading Ability: Evidence for Differential-Susceptibility Theory

**DOI:** 10.1101/2023.04.28.23289214

**Authors:** Qing Yang, Chen Cheng, Zhengjun Wang, Jay Belsky, Jingjing Zhao

**Author notes:** Address correspondence to: Jingjing Zhao; School of Psychology, Shaanxi Normal University 199 South Chang’an Road, Xi’an, China, 710062. Co-first author, equal contribution (Qing Yang, Chen Cheng).

## Abstract

While genetic and environmental factors have been shown as predictors of children’s reading ability, the interaction effects of identified genetic risk susceptibility and specified environmental for reading ability have rarely been investigated. The current study assessed potential gene-environmental (G×E) interactions on reading ability in 1477 school-aged children. The gene-environment interactions on character recognition were investigated by an exploration analysis between the risk single-nucleotide polymorphisms (SNPs) which were discovered by previous genome-wide association studies of developmental dyslexia (DD), and parental education (PE). The re-parameterized regression analysis suggested that this G×E interaction conformed to the strong differential-susceptibility model. Results showed that rs281238 exhibits a significant interaction with PE on character recognition. Children with “T” genotype profited from high PE, whereas they performed worse in low PE environment, but “CC” genotype children were not malleable in different PE environments.

**Research Highlights:**

- We calculated the Cumulative Genetic Score (CGS) of 9 SNPs related to developmental dyslexia, and found that the interaction between CGS and parental education on reading ability. The G×E comforted to the differential-susceptibility model that those individuals carrying more plasticity alleles were affected more than those carrying more fewer.
- The interaction between rs281238 and parental education conformed to the strong differential-susceptibility model that children with “T” allele would have highest reading ability in positive environment and lowest reading ability in adversity environment, whereas children without “T” allele would not be affected by parental education.

## Introduction

Developmental dyslexia (DD) is known to be a hereditary neurological disorder with unbalanced development between reading ability and age, which cannot be accounted for by low intelligence, inadequate education, visual or auditory acuity deficits or other mental disorders (Arbanas, 2015). The prevalence of DD is reported to be around 1.3-17.2% among school-age students varying in different orthographies and depending on the criteria used for diagnosis (Di Folco, Guez, Peyre, & Ramus, 2020) and its etiology is influenced by both genetic and environmental factors (Bishop, & Snowling, 2004).

Numerous studies have also suggested that environmental factors, such as birth weight, born preterm, home-literacy environment (HLE), socioeconomic status (SES) and parental education (PE), have important influences on children reading abilities’s development (Allotey et al., 2018; Aram, Korat, & Hassunah-Arafat, 2013; Fernald, Marchman, & Weisleder, 2013; Friend, DeFries, & Olson, 2008; Litt et al., 2012). The studies have highlighted the importance of family socioeconomic status in early childhood literacy development, and these findings consistently show that children from high socioeconomic status have higher reading and language skills before and after formal education than children from low socioeconomic status (Hoff, 2003;Noble, Farah, & McCandliss, 2006; Noble, McCandliss, & Farah, 2007; Rowe & Goldin-Meadow, 2009).

In recent years, a few studies have attempted to identify single nucleotide polymorphisms (SNPs) associated with reading or dyslexia by genome-wide association analysis study (GWAS) (Meaburn et al., 2008; Field et al., 2013; Gialluisi et al., 2014; 2019; 2020; Eicher et al., 2013; Truong et al., 2019). There were four variants genome-wide significantly associated with reading ability and dyslexia in the previous literature (Gialluisi et al., 2019). However, genome-wide association analysis ignored the influence of environment on gene expression, it is essential to consider the effect of environment in the variants.

At present, gene-environment interactions (G×E) on reading ability have been rarely studied, and the sample size of gene-environment interaction studies is usually four times larger than the sample size needed to discover the main effect of genes (Thomas, 2010). Therefore, we wanted to explore the feasibility of the interaction between the set of SNPs in GWAS and the environment.

Although research of G×E on reading ability is little, G×E on behavior have been well-studied in recent years (Manuck, & McCaffery, 2014; Rutter, 2006). For example, behavioral genetic studies with identical and fraternal twins have suggested that the degree of genetic influence on, or heritability of, individual differences in cognitive and reading abilities varies with SES (Turkheimer, Haley, Waldron, D’Onofrio, & Gottesman, 2003) or PE (Friend et al., 2008). The bioecological model is usually applied in twin studies that explain how does the heritability of certain phenotype vary with the environment. Friend et al. (2008) as the first twin study of G×E on reading disability found that genetic influence was higher among children whose parents had a high level of education, than those children whose parents had a lower level of education.

Consistently, molecular genetic studies further reported that dyslexic candidate genes’ influence on reading disability may depend on the environment. Mascheretti et al. (2013) explored five candidate genes’ (DYX1C1, DCDC2, ROBO1, KIAA0319, and TTRAP) interactions with a series of environmental factors. Results showed that children with DYX1C1 risk alleles would perform worse in adversity environment, such as low birth weight, low SES and history of smoking during pregnancy but would not be affected in positive environment. Su et al. (2015) investigated G×E on orthography processing in Chinese children and found that children carrying the minor allele of rs1091047 exhibited a smaller N170 effect in low home-literacy environment than those children from high hone-literacy environment. Both studies were consistent with the diathesis-stress model, which suggests that heritability for a particular behavior would be greater in poor environments while the deleterious gene would not be observed in more supportive environments and this model has been proposed to explain why certain behavioral disorders had a greater association with risk genes in environments where individuals have been exposed to a great deal of stressful life events (Caspi, et al., 2002; 2003).

Recently, however, a theoretical alternative to the diathesis-stress model has been proposed (i.e., the differential-susceptibility model, for summary, see Belsky & Pluess, 2013) and applied to the study of gene-environment interactions (Belsky & Pluess, 2009). The differential-susceptibility model framework stipulates that some individuals are not only more susceptible to negative environmental factors but also to positive ones as well. According to this model, some ‘risk genes’ might be better conceptualized as ‘plasticity genes’ (Belsky et al., 2009). In early literacy instruction area, Kegel, Bus, & IJzendoorn (2011) have found children had differential susceptibility of environment. Individuals with 7-repeat allele of the dopamine receptor D4 (DRD4-7R) profited most from the positive feedback of computer program, whereas they performed worst of early literacy skills in the absence of such feedback. And more recently, many other molecular genetic studies of gene-environment interactions have confirmed this method (Green et al., 2017; Wang, Tian, & Zhang, 2020).

So far, studies on G×E effect have mainly focused on Single Nucleotide Polymorphisms (SNPs), such as SNPs from previous genome-wide association (GWA) studies (Manuck & McCaffery, 2014). GWA study is a theoretical approache to gene discovery that is meant to find phenotype associated genetic variation without regard to biological pathway and function. Therefore, the 9 single-nucleotide polymorphisms (SNPs) from previous genome-wide association (GWA) studies in different cohorts in developmental dyslexia and reading ability were selected in the present study as genetic factors on reading ability of Chinese children. We focused on parental education (PE) as a marker of environmental variable. Parental education (PE) as an environmental factor has been shown to be a strong predictor for a variety of cognitive outcomes of children (Bradley & Corwyn, 2002; Craig, 2006). We predicted that if genetic and environmental influence were interdependent in reading ability, the effects of genetic signal should vary along the PE distribution. In confirmation analysis, two gene-environment interaction models (the diathesis-stress model vs. the differential-susceptibility model) were compared in a re-parameterized analysis to identify which model was a better fit to our data.

## Methods

### Participants

A total of 3217 primary school students from grade 3 to grade 6 were recruited from three provinces in northwestern part of China (Shaanxi Province, Gansu Province and Inner Mongolia Province). All these participants were school-aged children without any neurological disorder or neurotropic drugs history. In total, 2476 saliva samples of these participants were eligible for subsequently assaying genes and 2415 students’ genetic data were available at last. Finally, there were 1477 students had all the phenotype, genotype and environmental data (age = 116.34 ± 12.14 months, male/female = 737/740). Ethical approval was obtained from Shaanxi Normal University and written informed consent was obtained for all the participants’ parents.

### Measures

#### Genetic analyses

DNA was obtained from oral epithelial cells in students saliva samples and 2476 samples were genetyped using Illumina Asian screening array (650K) by Beijing Compass Biotechnology. Quality control was performed standard quality control metrics (Anderson et al., 2010; Chang, 2015) by using PLINK v1.9 (http://pngu.mgh.harvard.edu/purcell/plink/). Eight samples were excluded as they had sex discrepancies between the records and the genetically inferred data. Fifty-three samples were removed because had unexpected duplicates or probable relatives (PI-HAT>0.20). Next, SNPs were eliminated if they showed a variant call rate <0.95, a minor allele frequency (MAF) <0.02, a missing genotype data (mind) <0.90, or a hardy-weinberg equilibrium (HWE) <10^-5^ with each dataset. Then, autosomal variants were aligned to the 1000G genomes phase 1v3 reference panel for imputation, which follow the standard procedure consistent with previous GWAS studies (see Gialluisi et al., 2020). Finally, significant reading-related or dyslexia-related SNPs were extracted from our data.

We collected all the SNPs which were associated with reading and recorded by the GWAS Catalog (https://www.ebi.ac.uk/gwas/). There were 4 SNPs (rs1555839, rs17663182, rs349045 and rs16928927) showed a strict significant association with reading on genome-wide level (*p* ≤ 5 × 10^-8^), however, none of them existed in the current study sample. Thus, we loosened the criteria from *p* ≤ 5 × 10^-8^ to *p* ≤ 5 × 10^-7^. Then, there were 22 SNPs recorded by GWAS Catalog (for detailed information see Supplementary material Table S1) and 9 of them existed in our samples (see Table 1).

**Table 1.**
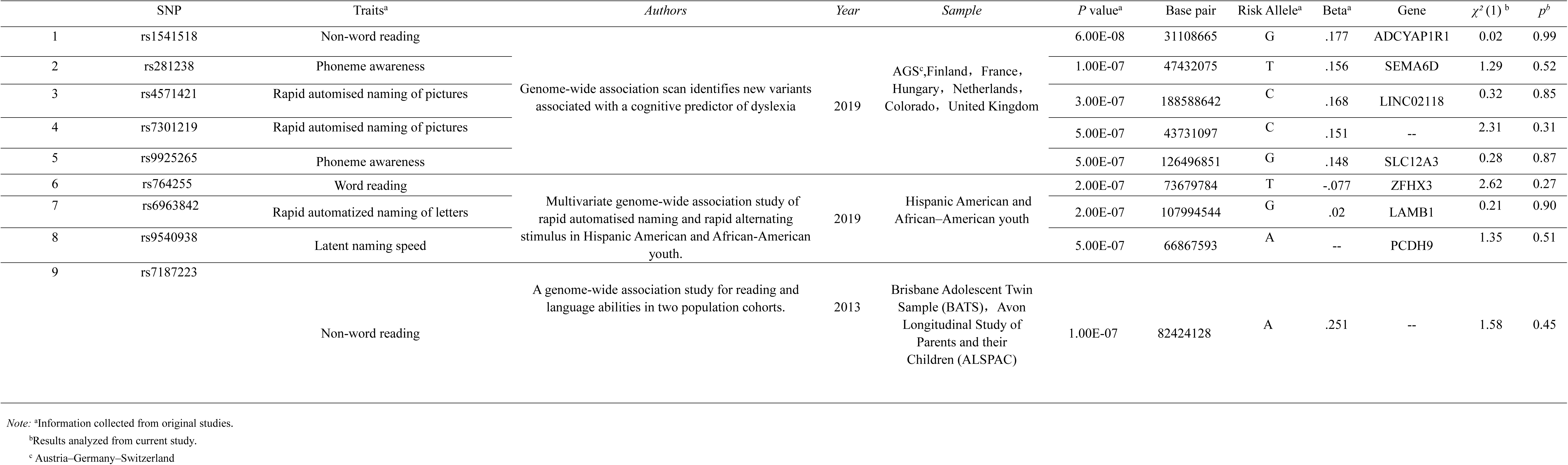
Detailed information of the nine selected SNPs.

| | SNP | Traits<sup>a</sup> | *Authors* | *Year* | *Sample* | *P* value<sup>a</sup> | Base pair | Risk Allele<sup>a</sup> | Beta<sup>a</sup> | Gene | $\chi^2(1)$<sup>b</sup> | $p^b$ |
| --- | --- | --- | --- | --- | --- | --- | --- | --- | --- | --- | --- | --- |
| 1 | rs1541518 | Non-word reading | Genome-wide association scan identifies new variants associated with a cognitive predictor of dyslexia | 2019 | AGS<sup>c</sup>,Finland, France, Hungary, Netherlands, Colorado, United Kingdom | 6.00E-08 | 31108665 | G | .177 | ADCYAP1R1 | 0.02 | 0.99 |
| 2 | rs281238 | Phoneme awareness |  |  |  | 1.00E-07 | 47432075 | T | .156 | SEMA6D | 1.29 | 0.52 |
| 3 | rs4571421 | Rapid automatised naming of pictures |  |  |  | 3.00E-07 | 188588642 | C | .168 | LINC02118 | 0.32 | 0.85 |
| 4 | rs7301219 | Rapid automatised naming of pictures |  |  |  | 5.00E-07 | 43731097 | C | .151 | -- | 2.31 | 0.31 |
| 5 | rs9925265 | Phoneme awareness |  |  |  | 5.00E-07 | 126496851 | G | .148 | SLC12A3 | 0.28 | 0.87 |
| 6 | rs764255 | Word reading | Multivariate genome-wide association study of rapid automatised naming and rapid alternating stimulus in Hispanic American and African-American youth. | 2019 | Hispanic American and African–American youth | 2.00E-07 | 73679784 | T | -.077 | ZFHX3 | 2.62 | 0.27 |
| 7 | rs6963842 | Rapid automatized naming of letters |  |  |  | 2.00E-07 | 107994544 | G | .02 | LAMB1 | 0.21 | 0.90 |
| 8 | rs9540938 | Latent naming speed |  |  |  | 5.00E-07 | 66867593 | A | -- | PCDH9 | 1.35 | 0.51 |
| 9 | rs7187223 | Non-word reading | A genome-wide association study for reading and language abilities in two population cohorts. | 2013 | Brisbane Adolescent Twin Sample (BATS), Avon Longitudinal Study of Parents and their Children (ALSPAC) | 1.00E-07 | 82424128 | A | .251 | -- | 1.58 | 0.45 |

*Note:* <sup>a</sup>Information collected from original studies.
<sup>b</sup>Results analyzed from current study.
<sup>c</sup> Austria–Germany–Switzerland

#### Reading ability

Each child’s reading ability was tested by C*hinese character recognition test*, a reading test used in Mainland China (Lei et al., 2011; Pan & Shu, 2014). The test consisted of 150 single Chinese characters selected from China’s *Elementary School Textbooks* (1996). The average frequency of the characters was 182 per million (ranging from 0 to 2282), and the reliability of this test was .95 (Pan & Shu, 2014). Each child was individually tested and required to read aloud each character at a time. Each child’s reading score, namely the number of characters reading correctly, was recorded. Finally, 2269 children completed this test.

#### Parental education (PE)

Students’ parental education (PE) were collected with questionnaire. Scores from 1 to 8 represent parents’ highest educational qualification from primary school, junior high school, senior high school, junior college, undergraduate, master, doctor to postdoctor, respectively. Both mother’s and father’s education level were collected. PE were the average of maternal and father’s highest educational achievement. In total, 1507 students’ PE information were obtained.

### Data analysis

Both exploratory and confirmatory analytic approaches consistent with Widaman et al. (2012) were used for 9 SNPs. Standard exploratory analysis was aimed to test whether there were G×E effects on character recognition. Confirmatory re-parameterized approach was employed to contrast the different hypothesis of G×E, i.e., strong and weak forms of the differential-susceptibility and diathesis-stress models to determine which provided the best, most parsimonious fit to the data.

#### Exploratory CGS

To examine the interaction between 9 SNPs and PE, we compute the cumulative-genetic socre (CGS), the risk allele was coded by 1 while the normal allele was coded by 0 (e.g. the two different alleles of rs281238 were C and T, and the risk allele was T, then genotype of CC, CT and TT was coded by 0, 1, 2, respectively). Parental education (PE) was the environmental variable *X_1_*, and a continuous variable CGS was the genetic variable *X_2_*. Age and sex were covariates.

The standard multiple regression model can be written as:

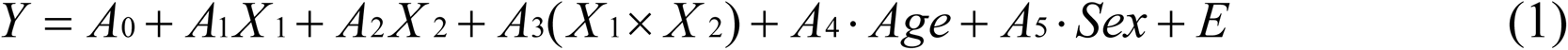

where *Y* is the dependent variable (reading ability); *A_0_* is the intercept, *A_1_* and *A_2_* are regression slopes for main effects of environment (*X_1_*) and CGS (*X_2_*), respectively; *A_3_* is the regression coefficient for the product variable (*X_1_ × X_2_*) and represents the difference in slope on *X_1_* for the CGS; *A_4_* and *A_5_* are the regression slopes for covariants age and sex; *E* is a stochastic error term.

#### Confirmatory CGS

Following Widaman et al. (2012), we re-parameterized the regression model, allowing a testing of alternative forms of the G×E interaction, as:

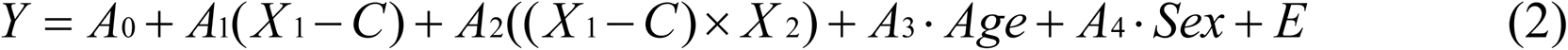

here Equation 2 is the re-parameterized the regression model for PRS and single SNP, respectively; *C* is the point on *X_1_ or X* at witch the slopes for the different PRS or genotype cross. If the cross point of *C* and its confidence interval (CI) is within the range of value on environment (*X_1_ or X*), the interaction tested is disordinal, reflecting differential-susceptibility model. On the contrary, if the cross point of *C* or its confidence interval (CI) is greater than or equal to the most positive point on environment (*X_1_ or X*) in this study, the interaction is ordinal, consistent with diathesis-stress model.

Next, to compare the efficiency of differential-susceptibility model and diathesis-stress model, we construct Model 3a, Model 3b. In Model 3a, if the cross point of *C* and its confidence interval (CI) is within the range of value on environment (*X_1_ or X*), the interaction tested is disordinal, reflecting differential-susceptibility model. On the contrary, if the cross point of *C* or its confidence interval (CI) is greater than or equal to the most positive point on environment (*X_1_ or X*) in this study, the interaction is ordinal, consistent with diathesis-stress model.

## Results

All the nine SNPs existed in our sample were conformed to Hardy Weinberg equilibrium(*p > .05,* see Table 1).

### Standard exploratory analysis

#### CGS

The standard multiple regression of CGS and PE without G×E was fit to data on character recognition. It had an *R^2^ = .2552* (*p < .001*), with environment effect significant, *Â1 = 1.87* (*SE = .43*), *p < .001*, but not the gene main effect, *Â2 = −.80* (*SE = .29*), *p = .78*. The main effects of covariates age and sex were also significant. Adding the G×E product term to the equation results in an increase in *ΔR^2^*, of .0023, as was the coefficient for the G×E product term itself, *Â3 = .52* (*SE = .24*), *p = .0307*. (See Table 2 and Figure 1). After ANOVA analysis, 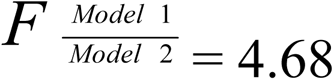, *F* ratio > 1.0 suggested that we can further to evaluate competing theoretical models (Belsky & Widaman, 2018).

**Table 2.** Results for alternative regression models for CGS on Character Recognition.

| Standard parameterization |  |  | Re-parameterized regression equation |  |  |
| --- | --- | --- | --- | --- | --- |
|  |  |  | Differential susceptibility |  | Diathesis-Stress |
|  | Gene(G) and environment(E) main effects: Model 1 | Main effects and G × E interaction: Model 2 | Parameter | Model 3a | Model 3b |
| $A_0$ | -7.05(6.01) | 5.69(8.41) | $C$ | 3.34(.56) | 6.85(--)<sup>a</sup> |
| $A_1$ | 1.87(.43) | -2.09(1.88) | $A_0$ | -1.28(5.02) | 5.06(4.59) |
| $A_2$ | -.80(.29) | -1.75(.82) | $A_1$ | -2.09(1.88) | 1.33(.72) |
| $A_3$ | - | .52(.24) | $A_2$ | .52(.24) | .07(.07) |
| $A_4$ | .92(.04) | .92(.04) | $A_3$ | 0.92(.04) | .92(.04) |
| $A_5$ | -2.56(.95) | -2.57(.95) | $A_4$ | -2.57(.95) | -2.573(0.95) |
| $R^2$ | .2552 | .2575 | $R^2$ | 0.2555 | 0.2536 |
| $F$ | 126.10 | 102.06 | $F$ | 127.70 | 126.40 |
| $df$ | 4,1472 | 5,1471 | $df$ | 5,1471 | 4,1472 |
| $p$ | <.0001 | <.0001 | $p$ | <.0001 | <.0001 |
| $F$ vs. 1 | -- | 4.68 | $F$ vs. 3b | 3.54 | -- |
| $df$ | -- | 1,1471 | $df$ | 1,1471 | -- |
| $p$ | -- | .0307 | $p$ | .0491 | -- |
| AIC | 122770.47 | 12767.78 | AIC | 12767.78 | 12769.67 |
| BIC | 12802.26 | 12804.87 | BIC | 12804.87 | 12801.46 |

Note: AIC, Akaike information criterion; BIC, Bayesian information criterion.
Tabled values are parameter estimates, with their standard errors in parentheses.
*F* vs. 1, 3a and 3c, stands for an *F* test of the difference in R<sup>2</sup> for Model 2 versus Model 1, Model 3b versus Model 3a, and Model 3d versus Model 3c, respectively.
<sup>a</sup>Parameter fixed at reported value; SE is not applicable, so is listed as (--).

**Figure 1.**
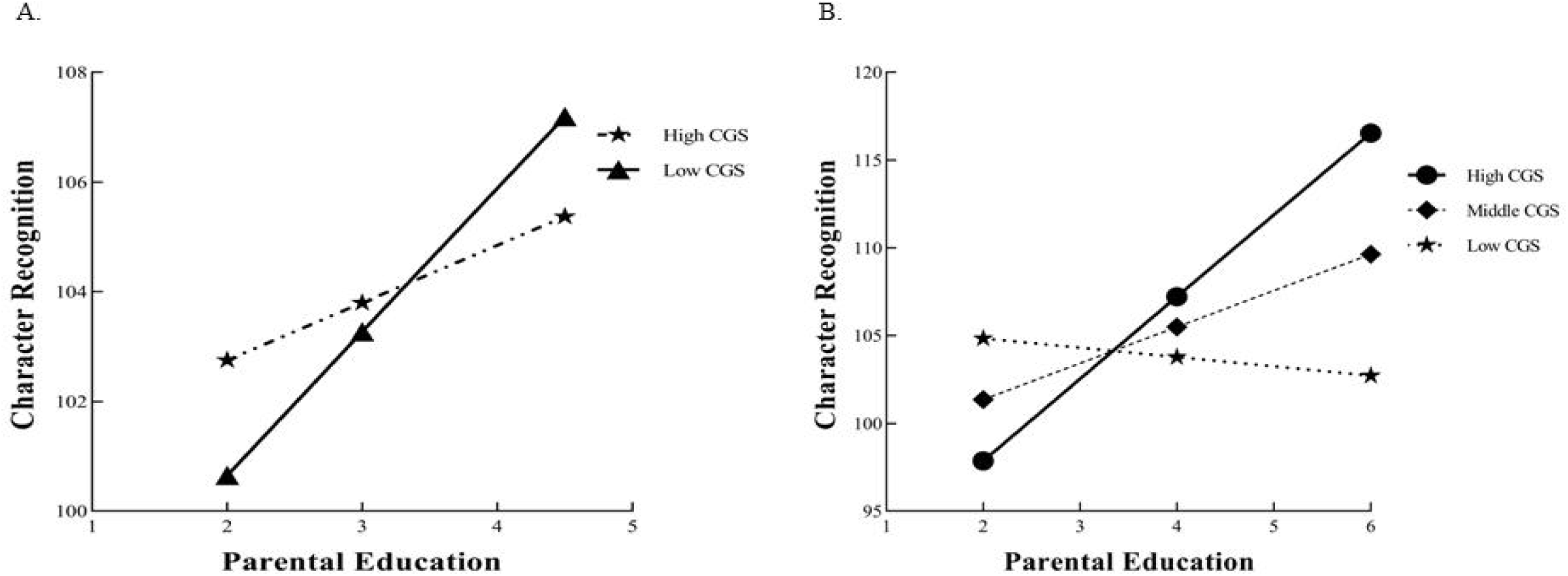
A. Simple slope analysis of Character Recognition in Low and High CGS subgroups; B. The plots for the results of the interaction between CGS and PE to predict Character Recognition in the differential susceptibility model.

#### Single SNP

After test the G×E of CGS, we verify the interactions of each single SNP (See Supplementary materials Section1, Table S2 and S3). Only the SNP of rs281238 was significant after Bonferroni correction as shown in Table S3. Results of the other SNPs are shown in supplementary materials Table S2. Model 1 in Table S3 has an *R^2^* = .2531 (*p* < .001), with a significant main effect of PE, *Â_1_ =* 1.88 (*SE* = .43), *p* < .001, but not the SNP main effect, *Â_2_ =* .01 (*SE* = .67), *p* = .98. The G×E interaction produced a significant Δ*R^2^* of .0035, *p* = .005 (see Model 2). The coefficients of rs281238, *Â_2_ =* −5.08 (*SE* = 1.94), *p* = .009, and G×E product term, *Â_3_ =* −1.56 (*SE* = .56), *p* = .005, and 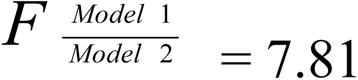, by ANOVA analysis, so confirmatory re-parameterized analysis can be estimated. Figure 2 shows the difference in the impact of environment on character recognition of different-genotypes.

**Figure 2.**
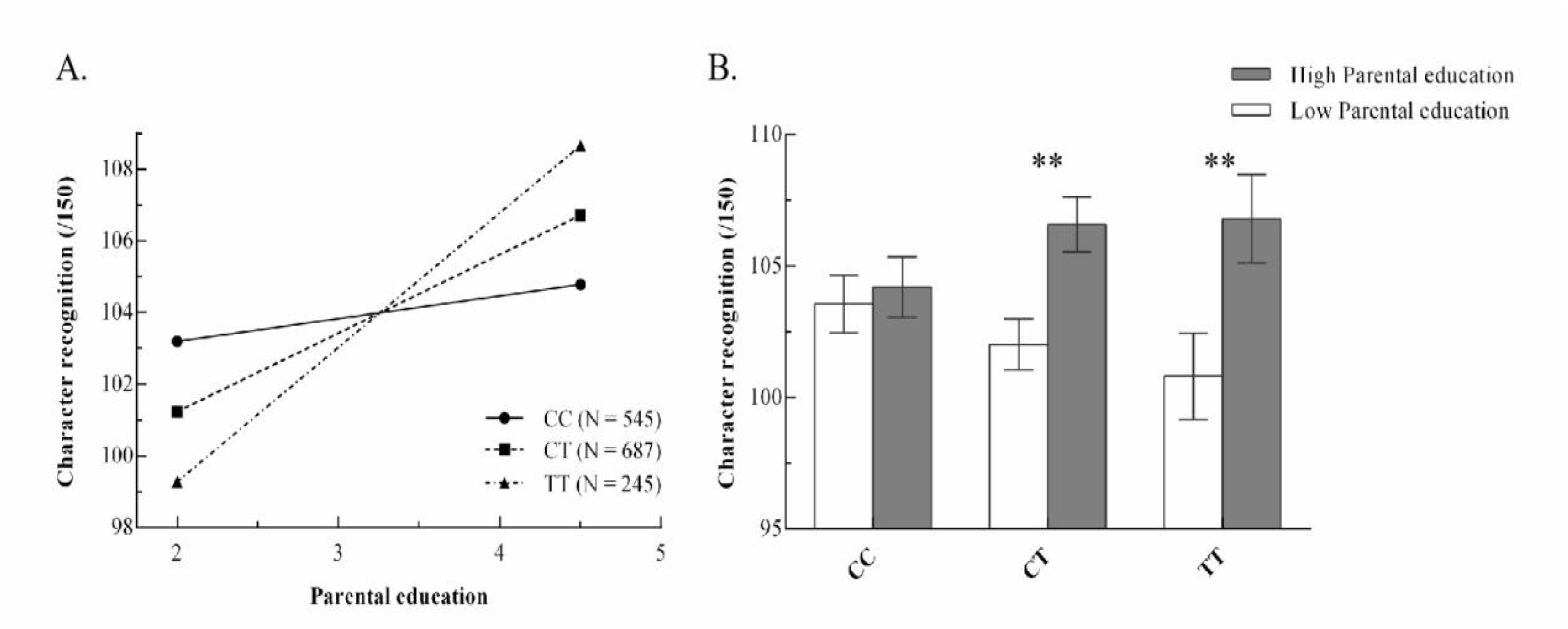
A. Simple slope analysis for character recognition in different genotype groups, and B. character recognition of different genotypes children in different environment (CC=279/266, CT=373/314, TT=128/117).

### Confirmatory re-parameterized analysis

Different versions of Equation 2 reflecting strong and weak forms of differential-susceptibility and diathesis-stress models for G×E interaction on character recognition.

#### Differential-susceptibility vs. diathesis-stress model for CGS

The estimated cross-over point *C* fell close to the sample mean on PE and the 95% CI of *C* fell within the range of PE. In Model 3a, *Ĉ =* 3.34 (*SE =* .56), the 95% CI of *Ĉ* [2.24, 4.43], and explained a significant amount of variance in character recognition, *R^2^* = .2555, *p* < .0001 (see Table 2). Choosing the highest PE value as *C* leads to Model 3b, the diathesis-stress model. In Model 3b, diathesis-stress model, explained a significant amount of variance in character recognition, *R^2^* = .2536, *p* < .0001. Compared with Model3b, Modle3a adds a parameter *Ĉ*, resulting in a significant increase in *R^2^*, *ΔR^2^ =* .0019*, p =* .049, so Model3a were accepted.

#### Differential-susceptibility vs. diathesis-stress model for CGS

The estimated cross-over point *C* fell close to the sample mean on PE and the 95% CI of *C* fell within the range of PE. In Model 3a, *Ĉ =* 3.21 (*SE =* .36), the 95% CI of *Ĉ* [2.50, 3.91], and in Model 3b, *Ĉ =* 3.21 (*SE =* .42), the 95% CI of *Ĉ* [2.39, 4.03]. Model 3a represent the strong differential-susceptibility model suggests that children without the risk allele T would be unaffected by PE, but children with different number of the risk allele T would be positively influenced by PE at different degrees. Model 3a explained a significant amount of variance in character recognition, *R^2^* = .2567, *p* < .0001 (See Section 2 & Table S3). The nested model of Model 3a, relaxing the constraint that *A_1_* = 0, leads to Model 3b, the weak differential-susceptibility model. Model 3b has a modest increase on explaining character recognition variance over Model 3a, but not reach the significant level, *ΔR^2^ =* .0002*, p* = .52. It suggested that adding a parameter *A_1_* in the model cannot increase the fitness of the model significantly. Thus, we accept the strong differential-susceptibility model and reject the weak differential-susceptibility model.

In Model 3c, the strong diathesis-stress model, children without risk allele T would not affect by PE, however, reading ability of children with risk allele T would increase with the improvement of PE, but never catch up with the children without allele T. Model 3c explained a significant amount of variance in character recognition, *R^2^* = .2452, *p* < .0001. The nested model of Model 3c, which relaxing the constraint that *A_1_* = 0, leads to Model 3d, the week diathesis-stress model which explained a large amount of additional variance over that explained by Model 3c, *ΔR^2^ =* .0078*, p =* .0001. Therefore, we found no significant basis for accepting the parsimonious Model 3c, putting support in favor of the weak diathesis-stress model as a more optimal representation of the data.

Since there are no statistical tests can evaluate the efficiency of Model 3a and Model 3d because of the same degree of freedom. Therefore, AIC and BIC values were employed to compare these two models. Since Model 3a has lower values in both AIC and BIC than Model 3d, we accepted Model 3a, the strong differential-susceptibility model as the best model for the current data. In order to provide an effective way to display the interactions in data, we plot predicted values under strong differential-susceptibility model. The plot of predicted character recognition score is shown in Figure S1, where the number of T allele determine children’s malleability. Below the cross-over point (low level of PE), children’s reading ability increase from low to high in TT group, TC group and CC group as a function of PE. However, above the cross-over point (high level of PE), TT and TC group shows consistently higher performance than CC group as a function of PE.

## Discussion

In the current study, we found that CGS and rs281238 had a significant interaction with parental education on Chinese children’s reading ability and this interaction effect was consistent with the differential-susceptibility model. To our knowledge, this is the first study that implemented a molecular genetic approach to investigate the interaction of environmental factor and SNPs associated with dyslexia from GWAS in reading ability by employing a multiple regression and re-parameterized analysis to distinguish gene-environment interaction between differential-susceptibility and diathesis-stress models.

We calculated cumulative gene scores for nine SNPs and found an interaction between CGS and parental education on reading ability, suggesting the effect of multiple genes on complex traits (Plomin & Davis, 2009). The study also demonstrated cumulative effects on genes (Tyler et al ., 2009; Steiger et al ., 2016), thus, those children carrying more plasticity alleles were influenced more than those carrying fewer on reading, providing new evidence for the differential-susceptibility model for G×E effect on reading ability. This finding also supplemented evidence supporting the differential-susceptibility model for G×E effect on behavioral traits (Belsky et al., 2013; Wang et al., 2020) which offers an alternative, evolutionary-inspired view: in order to improve adaptation of different environments, some individuals are simultaneously sensitive to both negative and positive experience (Bakermans-Kranenburg & van IJzendoorn, 2006). Although GWAS found significant loci, their explanatory rate for reading was very small. This finding supports the idea that G×E interaction as a possible aid in gene discovery(Thomas, 2010) and the consideration of environment in the GWAS studies might increase the efficiency of gene. Our results provide a good insight into the influence of environment as well as the presence of multiple genes.

Moreover, the current study was performed in normal school-age students and the reading ability is normally distributed, thus, these results might put an explanation for both children with high and low reading ability. Since positive environments can facility those children with risk alleles get a better reading performance, it is instructive for intervention plans. Future gene-environment research should consider both positive and adverse environmental factors.

More importantly, we observed that the gene-environment interactions between rs281238 and parental education on reading ability were consistent with differential-susceptibility model. The SNP, rs281238, locates in SEMA6D, a member of the semaphorin gene family, which codes a transmembrane protein that might play a navigational role in the signaling process of the nervous system (He, Wang, Koprivica, Ming, & Song, 2002). Other SNPs in SEMA6D, such as rs1378214, rs281320 and rs281323, have been reported a significant association (*p* < 5×10^-8^) with cognitive ability, attention deficit hyperactivity disorder (ADHD) and education attainment and have been replicated in independent studies (Kichaev et al., 2019; Klein et al., 2019; Lam et al., 2017; Lee et al., 2018; Okbay et al., 2016). SNPs in SEMA6D, such as rs12910916, rs2573570 and rs8039398 have also been reported related with reading, mathematics, cognition, education achievement and ADHD (Davies et al., 2018; Gialluisi et al., 2019; Lam et al., 2017; Lee et al., 2018; Martin et al., 2017; Rietveld et al., 2014). All these results suggest the impact of SEMA6D on cognitive and reading related traits. Taken together, our results put new evidence for the importance of SEMA6D in the complex cognition area - reading ability.

Notably, there are several limitations in the current study. First, genome-wide significant SNPs reported in previous literature of reading and dyslexia (Giallusisi et al., 2019) were not genotyped in our data and therefore were not tested for G×E effect on reading ability in this study. Future studies with genome sequencing of this Chinese cohort might be desirable to further test G×E effect on reading ability with genome-wide significant SNPs. Second, we only tested parental education as environmental factor. Other environmental factors might be valuable to be used to test G×E effect on reading ability in future studies. Finally, to further verify the external validity of this study, it is essential to replicate these findings in independent cohorts in future.

In summary, we have provided initial evidence for how the significant SNPs in developmental dyslexia GWA studies affect children’s reading performance by interacting with environmental factor of parental education. Our results indicated the validity of the differential-susceptibility model in reading ability.

## Data Availability

The beta values of the SNPs in this paper can be found in previous studies. The statistical methods and statistical data available from the corresponding author
upon reasonable request. The raw data are not publicly available due to legal or ethical restrictions.

## Acknowledgments

This work was funded by National Natural Science Foundation of China (61807023), Funds for Humanities and Social Sciences Research of the Ministry of Education (CN) (17XJC190010), Natural Science Foundation of Shaanxi Province (CN) (2018JQ8015), and Fundamental Research Funds for the Central Universities (CN) (GK201702011) to Jingjing Zhao. This study was also funded by Fund for Humanities and Social Sciences Research of the Ministry of Education (19YJC190023), and the China Postdoctoral Science Foundation funding project (2019M663924XB), Natural Science Foundation of Shaanxi Province (CN) (2021JQ-309), and Planning Subject for the 14th Five Year Plan of Shaanxi Education Sciences (SGH21Y0040) to Zhengjun Wang.

## Conflict of interest

The authors have no conflicts of interest to disclose.

## Data available statement

The beta values of the SNPs in this paper can be found in previous studies. The statistical methods and statistical data available from the corresponding author upon reasonable request. The raw data are not publicly available due to legal or ethical restrictions.

## Notes

### Competing Interest Statement

The authors have declared no competing interest.

### Author Declarations

Ethical approval was obtained from Shaanxi Normal University and written informed consent was obtained for all the participants' parents.

## Reference

Allotey, J., Zamora, J., Cheong-See, F., Kalidindi, M., Arroyo-Manzano, D., Asztalos, E., … Birtles, D. (2018). Cognitive, motor, behavioural and academic performances of children born preterm: a meta-analysis and systematic review involving 64 061 children. British Journal of Obstetrics and Gynaecology, 125(1), 16–25. doi:10.1111/1471-0528.14832

Anderson, C. A., Pettersson, F. H., Clarke, G. M., Cardon, L. R., Morris, A. P., & Zondervan, K. T. (2010). Data quality control in genetic case-control association studies. Nature Protocols, 5(9), 1564–1573 doi:10.1038/NPROT.2010.116

Aram, D., Korat, O., & Hassunah-Arafat, S. (2013). The contribution of early home literacy activities to first grade reading and writing achievements in Arabic. Reading and Writing, 26(9), 1517–1536. doi:10.1007/S11145-013-9430-Y

Arbanas, G. (2015). Diagnostic and statistical manual of mental disorders (DSM-5). In Codas (Vol. 51, pp. 61–64)

Bakermans-Kranenburg, M. J., & van IJzendoorn, M. H. (2006). Gene-environment interaction of the dopamine D4 receptor (DRD4) and observed maternal insensitivity predicting externalizing behavior in preschoolers. Developmental Psychobiology, 48(5), 406–409. doi:10.1002/DEV.20152

Belsky, J., & Pluess, M. (2009). Beyond Diathesis Stress: Differential Susceptibility to Environmental Influences. Psychological Bulletin, 135(6), 885–908. doi:10.1037/A0017376

Belsky, J., & Pluess, M. (2013). Beyond risk, resilience, and dysregulation: phenotypic plasticity and human development. Development and Psychopathology, 25, 1243–1261. doi:10.1017/S095457941300059X

Belsky, J., Pluess, M., & Widaman, K. F. (2013). Confirmatory and competitive evaluation of alternative gene-environment interaction hypotheses. Journal of Child Psychology and Psychiatry, 54(10), 1135–1143.

Belsky, J., & Widaman, K. (2018). Editorial Perspective: Integrating exploratory and competitive–confirmatory approaches to testing person × environment interactions. Journal of Child Psychology and Psychiatry, 59(3), 296–298.

Bishop, D. V. M., & Snowling, M. J. (2004). Developmental dyslexia and specific language impairment: same or different? Psychological Bulletin, 130(6), 858–886. doi:10.1037/0033-2909.130.6.858

Bradley, R. H., & Corwyn, R. F. (2002). Socioeconomic status and child development. Annual Review of Psychology, 53, 371–379. doi:10.1146/ANNUREV.PSYCH.53.100901.135233

Caspi, A., McClay, J., Moffitt, T. E., Mill, J., Martin, J., Craig, I. W., … Poulton, R. (2002). Role of Genotype in the Cycle of Violence in Maltreated Children. Science, 297(5582), 851–854. doi:10.1126/SCIENCE.1072290

Caspi, A., Sugden, K., Moffitt, T. E., Taylor, A., Craig, I. W., Harrington, H. L., … Braithwaite, A. (2003). Influence of Life Stress on Depression: Moderation by a Polymorphism in the 5-HTT Gene. Science, 301(5631), 386–389. doi:10.1126/SCIENCE.1083968

Chang, C. C., Chow, C. C., Tellier, L. C. A. M., Vattikuti, S., Purcell, S. M., & Lee, J. J. (2015). Second-generation PLINK: rising to the challenge of larger and richer datasets. GigaScience, 4(1), 7–7. doi:10.1186/S13742-015-0047-8v

Craig, L. (2006). Parental education, time in paid work and time with children: An Australian time-diary analysis. British Journal of Sociology, 57, 553–575. doi:10.1111/J.1468-4446.2006.00125.X

Davies, G., Lam, M., Harris, S. E., Trampush, J. W., Luciano, M., Hill, W. D., … Fawns-Ritchie, C. (2018). Study of 300,486 individuals identifies 148 independent genetic loci influencing general cognitive function. Nature Communications, 9(1), 2098–2098. doi:10.1038/S41467-018-04362-X

Di Folco, C., Guez, A., Peyre, H., & Ramus, F. (2020). Epidemiology of developmental dyslexia: A comparison of DSM-5 and ICD-11 criteria. medRxiv.

Eicher, J. D., Powers, N. R., Miller, L. L., Akshoomoff, N., Amaral, D. G., Bloss, C. S., … Casey, B. J. (2013). Genome-wide association study of shared components of reading disability and language impairment. *Genes*, Brain and Behavior, 12(8), 792–801. doi:10.1111/GBB.12085

Fernald, A., Marchman, V. A., & Weisleder, A. (2013). SES differences in language processing skill and vocabulary are evident at 18 months. Developmental Science, 16(2), 234–248. doi:10.1111/DESC.12019

Field, L. L., Shumansky, K., Ryan, J., Truong, D., Swiergala, E., & Kaplan, B. J. (2013). Dense-map genome scan for dyslexia supports loci at 4q13, 16p12, 17q22; suggests novel locus at 7q36. Genes, Brain and Behavior, 12(1), 56–69. doi:10.1111/GBB.12003

Fisher, S. E., & DeFries, J. C. (2002). Developmental dyslexia: genetic dissection of a complex cognitive trait. Nature Reviews Neuroscience, 3(10), 767–780.

Friend, A., DeFries, J. C., & Olson, R. K. (2008). Parental Education Moderates Genetic Influences on Reading Disability. Psychological Science, 19(11), 1124–1130. doi:10.1111/J.1467-9280.2008.02213.X

Gialluisi, A., Andlauer, T. F. M., Mirza-Schreiber, N., Moll, K., Becker, J., Hoffmann, P., … Brandler, W. (2019). Genome-wide association scan identifies new variants associated with a cognitive predictor of dyslexia. Translational Psychiatry, 9(1), 77. doi:10.1038/S41398-019-0402-0

Gialluisi, A., Andlauer, T. F. M., Mirza-Schreiber, N., Moll, K., Becker, J., Hoffmann, P., … Honbolygó, F. (2020). Genome-wide association study reveals new insights into the heritability and genetic correlates of developmental dyslexia. Molecular Psychiatry, 1–14. doi:10.1038/S41380-020-00898-X

Gialluisi, A., Newbury, D. F., Wilcutt, E. G., Olson, R. K., DeFries, J. C., Brandler, W. M., … Simpson, N. H. (2014). Genome-wide screening for DNA variants associated with reading and language traits. *Genes*, Brain and Behavior, 13(7), 686–701. doi:10.1111/GBB.12158

Green, C. G., Babineau, V., Jolicoeur-Martineau, A., Bouvette-Turcot, A.-A., Minde, K., Sassi, R., … Kennedy, J. L. (2017). Prenatal maternal depression and child serotonin transporter linked polymorphic region (5-HTTLPR) and dopamine receptor D4 (DRD4) genotype predict negative emotionality from 3 to 36 months. Development and Psychopathology, 29(3), 901–917. doi:10.1017/S0954579416000560

He, Z., Wang, K. C., Koprivica, V., Ming, G., & Song, H. J. (2002). Knowing How to Navigate: Mechanisms of Semaphorin Signaling in the Nervous System. Science Signaling, 2002(119). doi:10.1126/STKE.2002.119.RE1

Hoff, E. (2003). The specificity of environmental influence: Socioeconomic status affects early vocabulary development via maternal speech. Child Development, 74(5), 1368–1378.

Kegel, C. A. T., Bus, A. G., & IJzendoorn, M. H. van. (2011). Differential Susceptibility in Early Literacy Instruction Through Computer Games: The Role of the Dopamine D4 Receptor Gene (DRD4). *Mind*, Brain, and Education, 5(2), 71–78. doi:10.1111/J.1751-228X.2011.01112.X

Kichaev, G., Bhatia, G., Loh, P. R., Gazal, S., Burch, K., Freund, M. K., … Price, A. L. (2019). Leveraging Polygenic Functional Enrichment to Improve GWAS Power. American Journal of Human Genetics, 104(1), 65–75. doi:10.1016/J.AJHG.2018.11.008

Klein, M., Walters, R. K., Demontis, D., Stein, J. L., Hibar, D. P., Adams, H. H., … Sonuga-Barke, E. (2019). Genetic Markers of ADHD-Related Variations in Intracranial Volume. American Journal of Psychiatry, 176(3), 228–238. doi:10.1176/APPI.AJP.2018.18020149

Lam, M., Trampush, J. W., Yu, J., Knowles, E., Davies, G., Liewald, D. C., … Sundet, K. (2017). Large-Scale Cognitive GWAS Meta-Analysis Reveals Tissue-Specific Neural Expression and Potential Nootropic Drug Targets. Cell Reports, 21(9), 2597–2613. doi:10.1016/J.CELREP.2017.11.028

Lee, J. J., Wedow, R., Okbay, A., Kong, E., Maghzian, O., Zacher, M., … Linnér, R. K. (2018). Gene discovery and polygenic prediction from a genome-wide association study of educational attainment in 1.1 million individuals. Nature Genetics, 50(8), 1112–1121. doi:10.1038/S41588-018-0147-3

Litt, J. S., Taylor, H. G., Margevicius, S., Schluchter, M., Andreias, L., & Hack, M. (2012). Academic achievement of adolescents born with extremely low birth weight. Acta Paediatrica, 101(12), 1240–1245. doi:10.1111/J.1651-2227.2012.02790.X

Luciano, M., Evans, D. M., Hansell, N. K., Medland, S. E., Montgomery, G. W., Martin, N. G., … Bates, T. C. (2013). A genome-wide association study for reading and language abilities in two population cohorts. *Genes*, Brain and Behavior, 12(6), 645–652. doi:10.1111/GBB.12053

Manuck, S. B., & McCaffery, J. M. (2014). Gene-Environment Interaction. Annual Review of Psychology, 65(1), 41–70. doi:10.1146/ANNUREV-PSYCH-010213-115100

Martin, J., Walters, R. K., Demontis, D., Mattheisen, M., Lee, S. H., Robinson, E., … Lichtenstein, P. (2017). A Genetic Investigation of Sex Bias in the Prevalence of Attention-Deficit/Hyperactivity Disorder. Biological Psychiatry, 83(12), 1044–1053. doi:10.1016/J.BIOPSYCH.2017.11.026

Mascheretti, S., Bureau, A., Battaglia, M., Simone, D., Quadrelli, E., Croteau, J., … Maziade, M. (2013). An assessment of gene-by-environment interactions in developmental dyslexia-related phenotypes. *Genes*, Brain and Behavior, 12(1), 47–55. doi:10.1111/GBB.12000

Newbury, D. F., Paracchini, S., Scerri, T. S., Winchester, L., Addis, L., Richardson, A. J., Walter, J., Stein, J. F., Talcott, J. B., & Monaco, A. P. (2011). Investigation of dyslexia and SLI risk variants in reading- and language-impaired subjects. Behavior Genetics, 41(1), 90–104.

Noble, K. G., Farah, M. J., & McCandliss, B. D. (2006). Socioeconomic background modulates cognition–achievement relationships in reading. Cognitive Development, 21(3), 349–368.

Meaburn, E. L., Harlaar, N., Craig, I. W., Schalkwyk, L. C., & Plomin, R.. (2008). Quantitative trait locus association scan of early reading disability and ability using pooled dna and 100k snp microarrays in a sample of 5760 children. Molecular Psychiatry, 13(7), 729–740. doi:10.1038/SJ.MP.4002063

Okbay, A., Beauchamp, J. P., Fontana, M. A., Lee, J. J., Pers, T. H., Rietveld, C. A., … Meddens, S. F. W. (2016). Genome-wide association study identifies 74 loci associated with educational attainment. Nature, 533(7604), 539–542. doi:10.1038/NATURE17671

Plomin, R., Haworth, C. M., & Davis, O. S. (2009). Common disorders are quantitative traits. Nature Reviews. Genetics, 10(12), 872–878.

Price, K. M., Wigg, K. G., Feng, Y., Blokland, K., Wilkinson, M., He, G., … Lovett, M. W. (2020). Genome-Wide Association Study of Word Reading: Overlap with Risk Genes for Neurodevelopmental Disorders. *Genes*, Brain and Behavior, 19(6). doi:10.1111/GBB.12648

Raskind, W. H., Peter, B., Richards, T., Eckert, M. M., & Berninger, V. W. (2013). The genetics of reading disabilities: from phenotypes to candidate genes. Frontiers in psychology, 3, 601.

Rietveld, C. A., Esko, T., Davies, G., Pers, T. H., Turley, P., Benyamin, B., … Lee, J. J. (2014). Common genetic variants associated with cognitive performance identified using the proxy-phenotype method. Proceedings of the National Academy of Sciences of the United States of America, 111(38), 13790–13794. doi:10.1073/PNAS.1404623111

Rowe, M. L., & Goldin-Meadow, S. (2009). Differences in early gesture explain SES disparities in child vocabulary size at school entry. Science, 323(5916), 951–953.

Rutter, M. (2006). Genes and behavior: Nature-nurture interplay explained. *Malden*, MA: Blackwell.

Steiger, H., Thaler, L., Gauvin, L., Joober, R., Labbe, A., Israel, M., & Kucer, A. (2016). Epistatic interactions involving DRD2, DRD4, and COMT polymorphisms and risk of substance abuse in women with binge-purge eating disturbances. Journal of Psychiatric Research, 77, 8–14.

Su, M., Wang, J., Maurer, U., Zhang, Y., Li, J., McBride, C., … Shu, H. (2015). Gene-environment interaction on neural mechanisms of orthographic processing in Chinese children. Journal of Neurolinguistics, 33, 172–186. doi:10.1016/J.JNEUROLING.2014.09.007

Thomas, D. (2010). Gene–environment-wide association studies: emerging approaches. Nature Reviews Genetics,11(4), 259–272.

Truong, D. T., Adams, A. K., Paniagua, S., Frijters, J. C., Boada, R., Hill, D. E., … Wolf, M. (2019). Multivariate genome-wide association study of rapid automatised naming and rapid alternating stimulus in Hispanic American and African-American youth. Journal of Medical Genetics, 56(8), 557–566. doi:10.1136/JMEDGENET-2018-105874

Turkheimer, E., Haley, A., Waldron, M., D’Onofrio, B., & Gottesman, I. I. (2003). Socioeconomic status modifies heritability of IQ in young children. Psychological Science, 14(6), 623–628.

Tyler, A. L., Asselbergs, F. W., Williams, S. M., & Moore, J. H. (2009). Shadows of complexity: what biological networks reveal about epistasis and pleiotropy. BioEssays : news and reviews in molecular, cellular and developmental biology, 31(2), 220–227.

Wang, Tian, & Zhang, (2020). Interactions between the combined genotypes of 5-HTTLPR and BDNF Val66Met polymorphisms and parenting on adolescent depressive symptoms: A three-year longitudinal study. Journal of Affective Disorders, 265, 104–111. doi:10.1016/j.jad.2020.01.064

Widaman, K. F., Helm, J. L., Castro-Schilo, L., Pluess, M., Stallings, M. C., & Belsky, J. (2012). Distinguishing ordinal and disordinal interactions. Psychological Methods, 17(4), 615–622. doi:10.1037/A0030003

