## Supplementary materials for "Interaction between risk SNPs of Developmental Dyslexia and Parental Education on Reading Ability: Evidence for Differential-Susceptibility Theory"

Contents

Section1: Exploratory single SNP	2

Section2: Confirmatory single SNP	3

Table S1. Detailed information of the nine selected SNPs	5

Table S2. Results of the other eight SNPs multiple regression on character recognition	6

Table S3. Results for alternative regression models for rs281238 on reading ability	7

Figure S1. Plot of predicted values as a function of parental education under the strong differential-susceptibility gene-environment model for character recognition	8

**Section1: Exploratory single SNP**

Beyond the interaction of PRS and environment, we further test the gene-environment interactions for each of the nine SNPs with environment. The standard multiple regression model is similar with Equation 1:

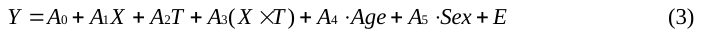

where *Y* is the dependent variable (reading ability, i.e., characters recognition scores); *X* is the environment variable (PE); while *T* is a trivariate (0 represented none risk alleles, 1 represented 1 risk allele and 2 represented 2 risk alleles); *X × T* is the product variable of gene-environment interaction; *A_1_* and *A_2_* are regression slopes for main effects of environment (*X*) and SNP (*T*), respectively; *A_3_* is the regression coefficient for the product variable (*X × T*) and represents the difference in slop on *X* among the “zero risk allele” group, “one risk allele group” and “double risk alleles group”; *A_0_* is the intercept, *A_4_* and *A_5_* are the regression slopes for covariant age and sex, respectively; *E* is a stochastic error term.

Equation 3 is fit once excluding the product term (*X × T*), testing main effects of SNP and PE (Model 1). The significant increase in the squared multiple correlation, *R^2^*, after adding the product term, provides evidence for G×E interaction (Model 2).

**Section2: Confirmatory single SNP**

Following Widaman et al. (2012), we re-parameterized the regression model, allowing a testing of alternative forms of the G×E interaction, as :

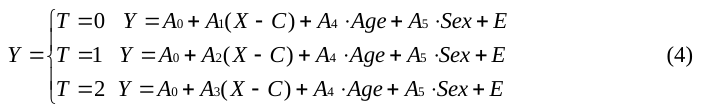

here Equation 4 is the re-parameterized the regression model for Equation 1; *C* is the point on *X* at which the slopes for the different genotype groups cross. If the cross point of *C* and its confidence interval (CI) is within the range of value on environment, the interaction tested is disordinal, reflecting differential-susceptibility model. On the contrary, if the cross point of *C* or its confidence interval (CI) is greater than or equal to the most positive point on environment in this study, the interaction is ordinal, consistent with diathesis-stress model.

Next, to compare the efficiency of strong or weak differential-susceptibility model and diathesis-stress model, we construct Model 3a, Model 3b, Model 3c and Model 3d. In Model 3a and Model 3b, we assume the cross-over point *C* fell at the range of *X* observed values and the G×E is disordinal. If the slope of none risk allele genotype group (*T* =0) is fixed at zero (i.e., *A_1_* = 0), the model (Model 3a) in Equation 2 is consistent with strong differential-susceptibility, which means that none risk allele group is unaffected by the environment. Relaxing the constraint that *A_1_* = 0 lead to the Model 3b, the weak differential-susceptibility model, which means the slope for the none risk allele genotype group differs significantly from zero. If we add a free parameter in Equation 2 can explain significantly more variance than Model 3a, we would accept Model 3b, otherwise we would accept Model 3a.

For Model 3c and Model 3d, we assume that *C* is the max(*X*), namely *Mean(X) +3 SD*. If *A_1_* is fixed and environment have no effect on none risk allele group, the model would conform to strong diathesis-stress model, Model 3c. If the none risk allele genotype group is affected by environment (i.e., *A_1_* ≠ 0), the model would reflect the week diathesis-stress model, the Model 3d. We would reject Model 3d, if there is no significant increase in *R^2^* compared with Model 3c.

Then, ANOVA (Analysis of Variance), the Akaike information criterion (AIC) and the Bayesian information criterion (BIC) are employed to evaluate the applicability between these four models. For both the AIC and BIC, the lower of the value, the more efficient of the model

**Table S1. Detailed information of the nine selected SNPs.**

|  | SNP | Traits^a^ | *P* value^a^ | Base pair | Risk Allele^a^ | Gene | *χ²* (1) ^b^ | *p^b^* |
| --- | --- | --- | --- | --- | --- | --- | --- | --- |
| 1 | rs281238 | Phoneme awareness | 1.00E-07 | 47432075 | T | SEMA6D | 1.29 | 0.52 |
| 2 | rs10010152 | Word reading | 7.00E-07 | 188588642 | G | LINC01060 | 1.93 | 0.38 |
| 3 | rs764255 | Word reading | 2.00E-07 | 73679784 | T | ZFHX3 | 2.62 | 0.27 |
| 4 | rs1687482 | Rapid automatized naming of letters | 7.00E-07 | 126496851 | T | UROC1 | 0.23 | 0.89 |
| 5 | rs1541518 | Non-word reading | 6.00E-08 | 31108665 | G | ADCYAP1R1 | 0.02 | 0.99 |
| 6 | rs6963842 | Rapid automatized naming of letters | 2.00E-07 | 107994544 | G | LAMB1 | 0.21 | 0.90 |
| 7 | rs8188533 | Latent naming speed | 6.00E-07 | 43731097 | T | -- | 0.69 | 0.71 |
| 8 | rs9540938 | Latent naming speed | 5.00E-07 | 66867593 | A | PCDH9 | 1.35 | 0.51 |
| 9 | rs7187223 | Non-word reading | 1.00E-07 | 82424128 | A | -- | 1.58 | 0.45 |

*Note:* ^a^Information collected from original studies. ^b^Results analyzed from current study.

**Table S2. Results of the other eight SNPs multiple regression on character recognition.**

| Standard parameterization: Main effects and G×E interaction | | | | | | | | | | | | | | | | |
| --- | --- | --- | --- | --- | --- | --- | --- | --- | --- | --- | --- | --- | --- | --- | --- | --- |
| Parameter | B_0_ | *p* | B_1_ | *p* | B_2_ | *p* | B_3_ | *p* | B_4_ | *p* | B_5_ | *p* | R^2^ | *F* | *df* | *p* |
| rs1687482 | -8.42(6.37) | .19 | 2.18(.94) | .02 | .58(2.06) | .78 | -.22(.59) | .71 | .92(.04) | <.001 | -2.55(.95) | .007 | .2527 | 100.8 | 1471 | <.001 |
| rs1541518 | -7.56(6.36) | .23 | 1.89(1.10) | .06 | -.04(2.16) | .99 | -.01(.63) | .99 | .92(.04) | <.001 | -2.56(.95) | .007 | .2526 | 100.8 | 1471 | <.001 |
| rs6963842 | -7.00(5.80) | .23 | 1.61(.52) | .002 | -1.44(2.40) | .55 | .66(.70) | .35 | .92(.04) | <.001 | -2.58(.95) | .007 | .2534 | 101.2 | 1471 | <.001 |
| rs8188533 | -11.10(7.90) | .16 | 3.09(1.67) | .06 | 1.93(3.12) | .54 | -.68(.91) | .45 | .92(.04) | <.001 | -2.56(.95) | .007 | .2529 | 100.9 | 1471 | <.001 |
| rs9540938 | -11.88(10.26) | .25 | 2.60(2.41) | .28 | 2.16(4.46) | .63 | -.38(1.25) | .76 | .92(.04) | <.001 | -2.59(.95) | .006 | .2528 | 100.9 | 1471 | <.001 |
| rs7187223 | -7.75(5.70) | .17 | 1.88(.46) | <.001 | .85(3.52) | .81 | .06(1.04) | .95 | .92(.04) | <.001 | -2.55(.95) | .007 | .2530 | 101.0 | 1471 | <.001 |
| rs10010152 | -12.23(6.92) | .08 | 3.18(1.27) | .01 | 2.86(2.46) | .24 | -.81(.73) | .27 | .92(.04) | <.001 | -2.54(.95) | .007 | .2533 | 101.1 | 1471 | <.001 |
| rs764255 | -8.36(5.75) | .15 | 2.26(.52) | <.001 | 2.23(2.31) | .33 | -.82(.64) | .20 | .92(.04) | <.001 | -2.56(.95) | .007 | .2537 | 101.4 | 1471 | <.001 |

**Table S3. Results for alternative regression models for rs281238 on reading ability.**

| Standard parameterization | | |  | Re-parameterized regression equation | | | | | |
| --- | --- | --- | --- | --- | --- | --- | --- | --- | --- |
|  |  |  |  |  | Differential susceptibility | |  | Diathesis-Stress | |
| Parameter | Gene(G) and environment(E) main effects: Model 1 | Main effects and G×E interaction: Model 2 |  | Parameter | Strong: Model 3a | Weak: Model 3b |  | Strong: Model 3c | Weak: Model 3d |
| A_0_ | -7.61(5.67) | -3.71(5.83) |  | C | 3.21（.36） | 3.21（.41） |  | 6.85(--)^a^ | 6.85(--)^a^ |
| A_1_ | 1.88(.43) | .63(.62) |  | A_0_ | -.90（4.82） | -1.70（5.00） |  | 6.21（4.60） | 5.08（4.60） |
| A_2_ | .01(.67) | -5.08(1.94) |  | A_1_ | -- | .43（.67） |  | -- | 1.77（.46） |
| A_3_ | -- | 1.56(.56) |  | A_2_ | 2.49（.60） | 2.52（.60） |  | .48（.26） | 1.89（.45） |
| A_4_ | .92(.04) | .92(.04) |  | A_3_ | 3.27（.97） | 3.30（.97） |  | .68（.36） | 2.09（.52） |
| A_5_ | -2.56(.95) | -2.54(.95) |  | A_4_ | .91（.04） | .92（.04） |  | .86（.04） | .92(.04) |
| R^2^ | .2531 | .2566 |  | A_5_ | -2.53（.95） | -2.54（.95） |  | -2.48（.95） | -2.56（.95） |
| *F* | 126.1 | 102.9 |  | R^2^ | .2567 | .2569 |  | .2452 | .2530 |
| *df* | 4, 1472 | 5, 1471 |  | *F* | 102.90 | 103.00 |  | 120.90 | 101.00 |
| *p* | <.0001 | <.0001 |  | *df* | 5,1471 | 6,1470 |  | 4,1472 | 5,1471 |
| *F* vs. 1 | -- | 7.81 |  | *p* | <.0001 | <.0001 |  | <.0001 | <.0001 |
| *df* | -- | 5, 1471 |  | *F* vs. 3b | .41 | -- |  | 11.175 | 7.66 |
| *p* | -- | .0052 |  | *df* | 1,1470 | -- |  | 2,1470 | 1,1470 |
|  |  |  |  | *p* | .5202 | -- |  | <.0001 | .0057 |
|  |  |  |  | *F* vs. 3c | 21.95 | 11.75 |  | -- | 14.27 |
|  |  |  |  | *df* | 1,1471 | 2,1470 |  | -- | 1,1471 |
|  |  |  |  | *p* | <.0001 | <.0001 |  | -- | .0001 |
| AIC | 12770.55 | 12764.73 |  | AIC | 12764.51 | 12766.09 |  | 12784.38 | 12771.77 |
| BIC | 12802.34 | 12801.81 |  | BIC | 12801.59 | 12808.47 |  | 12816.17 | 12808.85 |

Note: AIC, Akaike information criterion; BIC, Bayesian information criterion.

Tabled values are parameter estimates, with their standard errors in parentheses.

*F* vs. 1, 3b and 3c, stands for an *F* test of the difference in R^2^ for Model 2 versus Model 1, Model 3a, 3c, 3d versus Model 3b, and Model 3a, 3b, 3d versus Model 3c, respectively.

^a^ Parameter fixed at reported value; SE is not applicable, so is listed as (--)

**Figure S1. Plots of predicted values as a function of parental education under the strong differential-susceptibility gene-environment model for character recognition.**

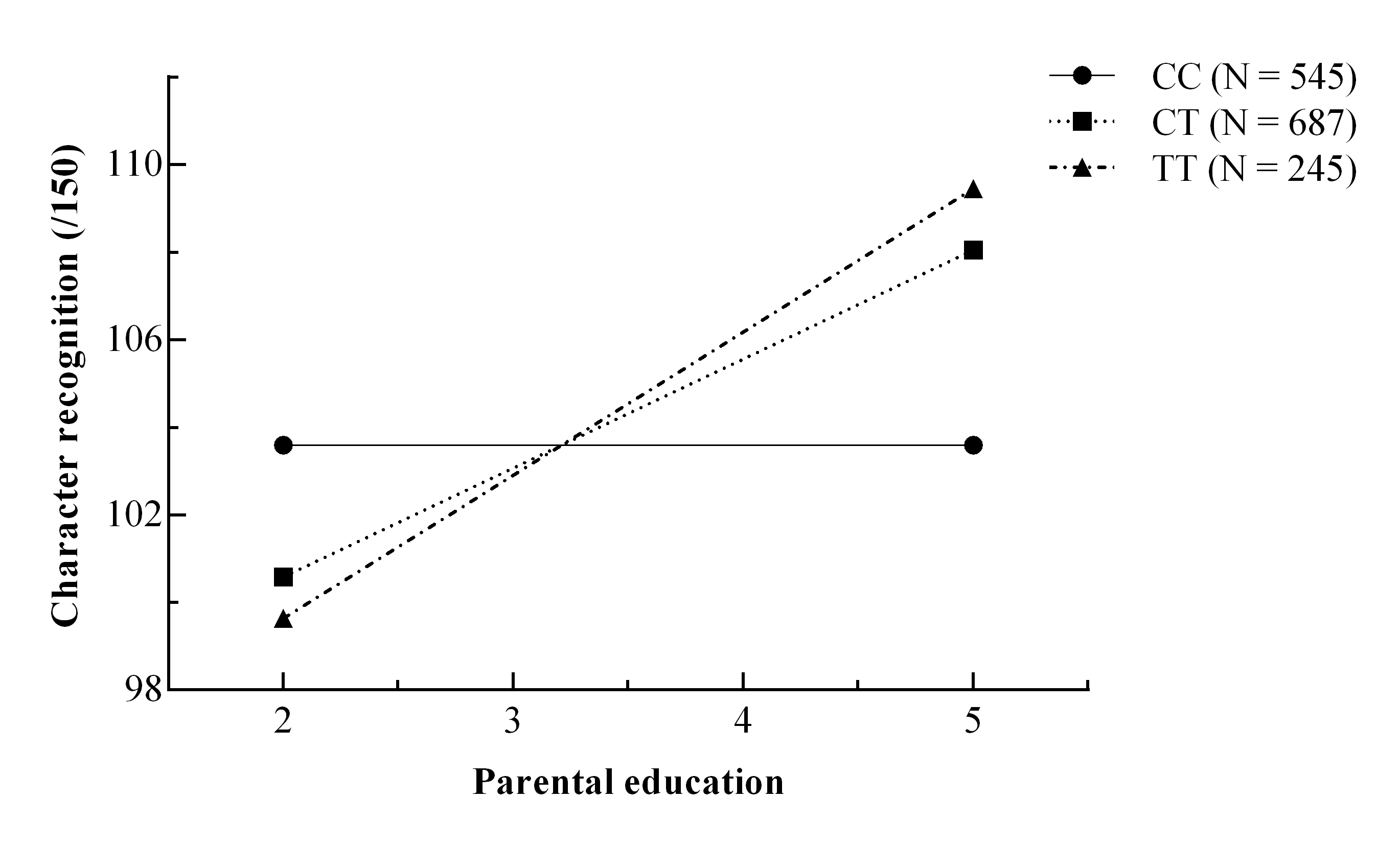
